# Distribution of non-falciparum malaria among symptomatic malaria patients in Dschang, West Region of Cameroon

**DOI:** 10.1101/2025.03.24.25324523

**Authors:** Pacome V. K. Tchuenkam, Samuel J. White, Varun Potlapalli, Eva M. Keming, Meredith S. Muller, Darlin B. N. Kaunda, Oksana Kharabora, Rhoel R. Dinglasan, Jonathan B. Parr, Christopher B. Tume, Jessica T. Lin, Jonathan J. Juliano, Innocent M. Ali

## Abstract

**Background:** Malaria is a vector-borne parasitic disease that continues to be a global public health threat. Five different species of the genus *Plasmodium* (*P. falciparum, P. malariae, P. ovale curtisi, P. ovale wallikeri*, and *P. vivax)* cause malaria in Sub-Saharan Africa. Previous cross-sectional surveys from 2013 and 2017 indicated the circulation of *P. vivax* in the West region of Cameroon, prompting an investigation into the prevalence of all non-falciparum malaria parasite species in this region.

**Methods:** A cross-sectional facility-based study recruited 431 clinically suspected cases of malaria from three health centres in the West region of Cameroon in 2020. Socio-demographic, clinical data, and dried blood spots (DBS) were collected from all consenting patients. Parasite DNA was extracted from DBS for real-time PCR amplification of species-specific *Plasmodium* 18S rRNA for *P. falciparum, P. ovale, P. malariae, and P. vivax*. In addition, *P. ovale* was further sub-classified into *P. ovale curtisi* or *P. ovale wallikeri*. The prevalence of different species was measured.

**Results:** Among the 431 samples, the overall malaria prevalence was 54.8% (236/431). Of these, 49.9% were infected with *P. falciparum*, 4% with *P. ovale*, and 0.9% with *P. malariae*. No *P. vivax* was detected. Mixed infections were common, with 3.5% of the infections harbouring more than one *Plasmodium* species. A total of 5 *P. ovale* and 1 *P. malariae* mono-infections were detected. Of the 17 *P. ovale* infections, 12 were successfully genotyped, with 6 *P. ovale curtisi*, 3 *P. ovale* wallikeri, and 3 mixture of the species.

**Conclusions:** While falciparum remains the dominant malaria parasite species among acute febrile illness cases, non-falciparum malaria is also commonly found in Dschang, both as a co-infection with *P. falciparum* and as mono-infections. Both subspecies of *P. ovale* are present in the region. Continued monitoring of non-falciparum species is needed for understanding malaria burden in West Cameroon.

## Introduction

Malaria is an infectious disease caused by parasites of the genus *Plasmodium* and transmitted by infected Anopheles female mosquitoes in tropical and subtropical countries (1). Despite substantial control efforts, it remains one of deadliest diseases in the world, with Africa suffering more than 95% of the burden. Several strategies are implemented across Africa to control malaria, including chemoprophylaxis, vector control, and appropriate diagnosis and treatment (2). High quality malaria surveillance is a key part of the WHO malaria containment strategy and is an important piece to investigate the impact of the disease on the health status of the population (3). These surveys should be used to determine the parasite species infecting individuals as optimal control measures and treatment varies from one species to another (3). To date, six species of malaria have been identified that cause disease in humans, *Plasmodium falciparum, P. malariae, P. vivax, P. knowlesi, P. ovale wallikeri* and *P. ovale curtisi*, of which 5 occur in Africa. *P. falciparum* and *P. vivax* are the most widely distributed world-wide with *P. falciparum* accounting for over 95% of the world’s cases and 99% of deaths from malaria. However, non-falciparum infections are increasingly recognized as sources of clinical morbidity and mortality (4).

Historically, it was thought that *P. vivax* was largely absent from West and Central Africa due to a majority of Sub-Saharan Africa population lacking the Duffy Antigen Receptor for Chemokines (DARC), the primary receptor for human red blood cell invasion (5). However, recent findings have reported an apparent increase in non-falciparum malarias in Cameroon, (6–8) and other regions of Africa (9–11). A study in Cameroon from 2017 found that 38.6% of all malaria cases in Dschang were *P. vivax* infections (6). This is almost twice (20.4%) the prevalence that Ngassa *et al*. reported in 2016 in Douala and far greater (4%) than reported by Fru-cho in 2014 (12). This inconsistency in the burden of *P. vivax* highlights the need to further characterise this species in West and Central Africa.

*P. ovale* is often detected as a mixed species infection in areas with high malaria transmission intensity, including Cameroon (13,14). The low parasite counts and mixed infections commonly results in misdiagnosis of *P. ovale* malaria, and its morphological similarities with *P. vivax* make microscopic identification challenging (15). Adding to the difficulty in species identification, there are two morphologically indistinguishable but genetically distinct species of *P. ovale*; *Plasmodium ovale curtisi* and *Plasmodium ovale wallikeri* (16). In Cameroon, *P. ovale curtisi* was first reported in Douala by Foko et al in 2021(17) in mixed infections with *P. falciparum*, but no occurrence of *P. ovale wallikeri* has been reported. According to Antonio-Nkondjio and colleagues, *P. malariae* represent 1% of infection cases in Cameroon by microscopy (18). This likely underrepresents the true burden as recent studies using molecular tools indicated 17% out of 236 blood samples analyzed (19) in a forest area and close to 20% (both in mono and mixed infections) in the Adamaoua region contained *P. malariae* (4). These studies suggest the need to deploy molecular diagnostics to improve non-falciparum species detection (18).

Cameroon primarily uses rapid diagnostic tests (RDT) that only detect *P. falciparum* antigens, providing no data on the burden of non-falciparum malaria (20,21). Diagnosis of non-falciparum infections are difficult, especially as current RDTs do not reliably or specifically detect *P. malariae, P. ovale spp*., and *P. vivax*. Also, the life cycles of *P. ovale spp*. and *P. vivax* include dormant liver stages, called hypnozoites, that can cause relapse weeks to months after primary infection and are not detectable by current diagnostics. With the aim to strengthen control measures and design specific tools to combat malaria, the main objective of this work was to characterize the burden of non-falciparum malaria in Dschang and surrounding neighborhoods by leveraging sensitive molecular diagnostics.

## Methods

### Ethics statement

The study was reviewed and approved by the institutional review board (IRB) of the Cameroon Baptist Convention Health Board (FWA00002077), Protocol IRB2019-40. Written informed consent was administered in French or English based on participant preference, via an independent translator (who also spoke the local language Yemba) as needed. For children, informed consent was obtained from a parent or guardian. Humans Subjects Research-exempt, de-identified dried blood spots were sent to the University of North Carolina for molecular testing.

### Study site and study design

The Dschang Health District encompasses 22 health areas and covers 1060 km^2^. It is a tropical, semi-urban environment at an elevation of 1380-1400m above sea level with a rainy season that occurs between mid-March–mid October. This study was a prospective hospital-based cross-sectional survey in the three main health facilities in the Dschang Health District in the West region of Cameroon: Dschang district hospital, St. Vincent Catholic Hospital, and “*Hopital des Soeur Servante du Christ de* Batseng’la”. This study was designed to characterize the species of parasites causing clinical malaria in this region of Cameroon.

### Patient enrolment and Sample collection

In total, 431 patients were enrolled between June 12, 2020–September 8, 2020 (Supplemental Table 1). The inclusion criteria of the study included fever (axillary temperature 37.5°C or self-reported history of fever) in past 24 hours without signs and symptoms suggestive of severe malaria. All those who signed the consent form were enrolled in the study. Participants were excluded if they had used anti-malarial medicine in the past 14 days. Dried blood spots (DBS) were prepared using left over blood samples by spotting 3 to 4 drops on a Whatman filter paper (N°03) and left to dry overnight away from sunlight. Samples were then sealed in a zip lock bag with a desiccant and stored at −20°C until DNA extraction.

**Table 1:** Presentation of socio-demographic data with respect to malaria outcome.

|  | **Total** | **Malaria Outcome** |  |  |
| --- | --- | --- | --- | --- |
|  |  | Malaria + | Malaria – | P-value |
| **Number of participants [n (%)]** | 431 (100) | 236 (54.8) | 195 (45.2) |  |
| ***Demographics*** |  |  |  |  |
| **Age (years)** |  |  |  |  |
| **<5** | 46 (10.7) | 28 (6.5) | 18 (4.2) | 0.17 |
| **6–14** | 27 (6.3) | 19 (4.4) | 8 (1.4) |  |
| **15–49** | 276 (64.0) | 142 (33.0) | 134 (31.1) |  |
| **>50** | 82 (19.0) | 41 (9.5) | 41 (9.5) |  |
| ***Gender*** |  |  |  |  |
| **Female** | 253 (58.7) | 125 (29.0) | 128 (29.7) | 0.06 |
| **Male** | 178 (41.3) | 105 (24.4) | 73 (17.0) |  |
| ***Level of Education*** |  |  |  |  |
| **Primary** | 159 (36.9) | 88 (15.6) | 71 (16.5) | 0.16 |
| **Secondary** | 58 (13.5) | 37 (8.6) | 21 (4.5) |  |
| **Tertiary** | 214 (49.6) | 105 (24.4) | 109 (25.3) |  |
| ***Presence of water source (within 2 min. walk)*** |  |  |  |  |
| **Yes** | 67 (15.6) | 41 (9.5) | 26 (6.0) | 0.16 |
| **No** | 355 (82.4) | 181 (42.0) | 174 (40.4) |  |
| **Do not know** | 9 (2.0) | 8 (1.9) | 1 (0.2) |  |
| ***Slept under a bednet the last 24 hours*** |  |  |  |  |
| **Yes** | 237 (55.0) | 130 (30.2) | 107 (24.8) | 0.44 |
| **No** | 182 (42.2) | 92 (21.4) | 90 (20.9) |  |
| **Did not respond** | 12 (2.8) | 8 (1.9) | 4 (0.9) |  |
| ***Keeps animals*** |  |  |  |  |
| **Yes** | 42 (09.7) | 21 (04.9) | 21 (4.9) | 0.87 |
| **No** | 379 (88.0) | 204 (47.3) | 175 (40.6) |  |
| **Did not respond** | 10 (2.3) | 5 (1.2) | 5 (1.2) |  |

### Detection of malaria parasites in clinical samples

One 6-mm whole punch of each DBS was used for molecular testing. Parasite DNA was extracted from DBS punch using Chelex®-100 (Bio-Rad Laboratories, CA, USA), extraction (22). Malaria positivity was assessed using quantitative real-time PCR (qPCR) for *P. falciparum, P. vivax, P. ovale spp*., and *P. malariae* using species-specific 18S rRNA gene fragment probes (Supplementary Table 1). All plates contained negative controls and positive controls consisting of mocked blood spots containing the desired target (MRA 177 178 179 and 180, BEI Resources, Manassas, VA). All reactions were carried out on a Bio-Rad CFX Connect Real-Time PCR Detection System. ROCHE FastStart Universal Probe Master mix (catalog #4913957001) was used alongside published primer sequences. Each reaction had a total volume of 10µl and was run for 45 cycles (Supplemental Table 1).

### Detection of *P. ovale wallikeri* and *curtisi species*

A combination of both nPCR (nested) and qPCR was used for the detection of *P. ovale spp*. as described by Potlapalli *et al*. (23). Briefly, the nPCR was performed using the specific primers rPLU1 and rPlU5 targeting a 1200bp gene fragment for nest 1 and the combination ROVA1 and ROVA2 for *P. ovale curtisi*; ROVA1v and ROVA2v for *P*.*ovale wallikeri*. For the qPCR, a modified protocol from Perandin *et al*. (24) and Calderaro *et al*, (25) targeting the small unit of the 18S rRNA was used. The reaction was carried out using Fast Start Universal Probe Master mix (ROX, Roche) and published primer and probe concentrations on a Bio-Rad CFX Connect Real-Time PCR Detection System. Both assays were run in parallel to 50 cycles, instead of the originally published 45 and 55 cycles. For *P. ovale curtisi* amplification, OVA-F (TTTTGAAGAATACATTAGGATACAATTAATG) and OVA-R (CATCGTTCCTCTAAGAAGCTTTACAAT) were used along with OVA (VIC) probe (CCTTTTCCCTATTCTACTTAATTCGCAATTCATG). For *P. ovale wallikeri* amplification, OVA-Fv (TTTTGAAG AATATATTAGGATACATTATAG) and OVA-R were used along with Ovav (FAM) probe (CCTTTTCCCTTTTCTACTTAATTCGCTATTATG). A common annealing temperature of 52.8°C was chosen as this yielded similar Ct thresholds for detection of the target plasmid copy concentrations (23).

### Statistical Analysis

Clinical and socio-demographic data were analyzed with Chi-square analysis for categorical variables. A Chi-square test was conducted to compare categorical variables to the outcome of interest. Fisher’s exact test was conducted for variables with less than 5 observations when appropriate. Bivariate associations between *Plasmodium* infection and socio-demographic data were investigated using R (4.4.1) studio.

## Results

### Socio-demographic characteristics and clinical parameters

Of the total participants enrolled, 59.3% (255/431) were from the District Hospital Dschang (HDD) and the rest enrolled from Hopital des Soeur servant du Christ de Batsengla (HSSCB, n=106 (24.6%)) and Hopital Saint Vincent (HSV, n=70 (16.2%)). Table 1 summarizes the key socio-demographic characteristics of the study population and association with *Plasmodium* infection status. In the study population, 58.1% (253/431) were female while 15/253 (5.9%) of the women were pregnant at the time of enrolment. The median age was 26 years (IQR: 20.7-31.3), and children less than 10 years represented 10% of the total study population. Close to 63% of the study population had either secondary or tertiary education. With regards to occupation, only 9% had a job in the formal sector. The majority were students (61%) as Dschang is a university town. While 86% reported travel out of the region (within Cameroon) in the past year at least once, most participants reported to have been in Dschang in the previous three months (77.9%). Most of the patients (82.3%) did not have a water source within 1km of their homes. About 10% of study participants reported keeping domestic animals at home. More than half of the study population (54.5%) reportedly owned bed nets. About the same proportion of all study participants reported sleeping under a bed net the previous night (55.2%). Of these socio-demographic data, only sex was associated with malaria positivity, with males more likely than females to test malaria positive with borderline significance (P=0.06).

Table 2 shows the symptoms at presentation. Among the 431 participants enrolled in this study, 311 reported symptoms beyond fever or history of fever the most commons symptoms were abdominal pain or diarrhea (35.7%), headache (29.5%), and general fatigue (28.3%). A minority of patients had cough, gastric pain, or other symptoms.

**Table 2.** Symptoms other than fever reported by participants.

|  | Malaria + | Malaria - | P.value |
| --- | --- | --- | --- |
| **Participants (n=431)** |  |  |  |
| **Headache (n=127; )** | 72 | 55 | 0.40 |
| **General fatigue (n=122)** | 66 | 56 | 0.88 |
| **Abdominal pain/diarrhoea (n=154)** | 79 | 75 | 0.52 |
| **Vomiting (n=22)** | 16 | 6 | 0.10 |
| **Cough (n=29)** | 16 | 13 | 1 |
| **Gastric pain (n=2)** | 2 | 0 | 0.54 |
| **Other (n= 25)** | 12 | 13 | 0.72 |
| **No symptoms reported other than fever or history of fever (n=118)** | 62 | 56 | 0.92 |

*This table represents symptoms other than fever or reported history of fever*

### Frequency of *Plasmodium* species, distribution, and associated symptomatology

Of the 431 patients enrolled, we observed an overall malaria prevalence by PCR of 54.8% (236/431). Infection status of malaria infected participants is summarized in Figure 2. Of these 431, we observed that 230/431 (53.36%) were positive for *P. falciparum*, 17/431 (4.0%) were positive for *Plasmodium ovale spp*. and 4/431 (0.9%) were infected with *P. malariae*. No cases of *Plasmodium vivax* were identified. The remaining 195 of 431 (45.2%) febrile illness samples were negative for any *Plasmodium* species by PCR. Mixed infections were not uncommon. Of all malaria-positive individuals (n=236), 8.9% (21/236) contained at least one non-falciparum species, with 7.2% (17/236) and 1.7% (4/236) containing *P. ovale* spp. and *P. malariae*, respectively. We observed that among 17 samples that were positive for *P. ovale*, 5/17 were mono-infections (29%) and 12/17 (71%) were mixed infections with *P. falciparum*. Whereas, only a single sample was a *P. malariae* mono-infection (1/4, 25%) and the remaining 3/4 (75%) were mixed infections with *P. falciparum*.

**Figure 1:**
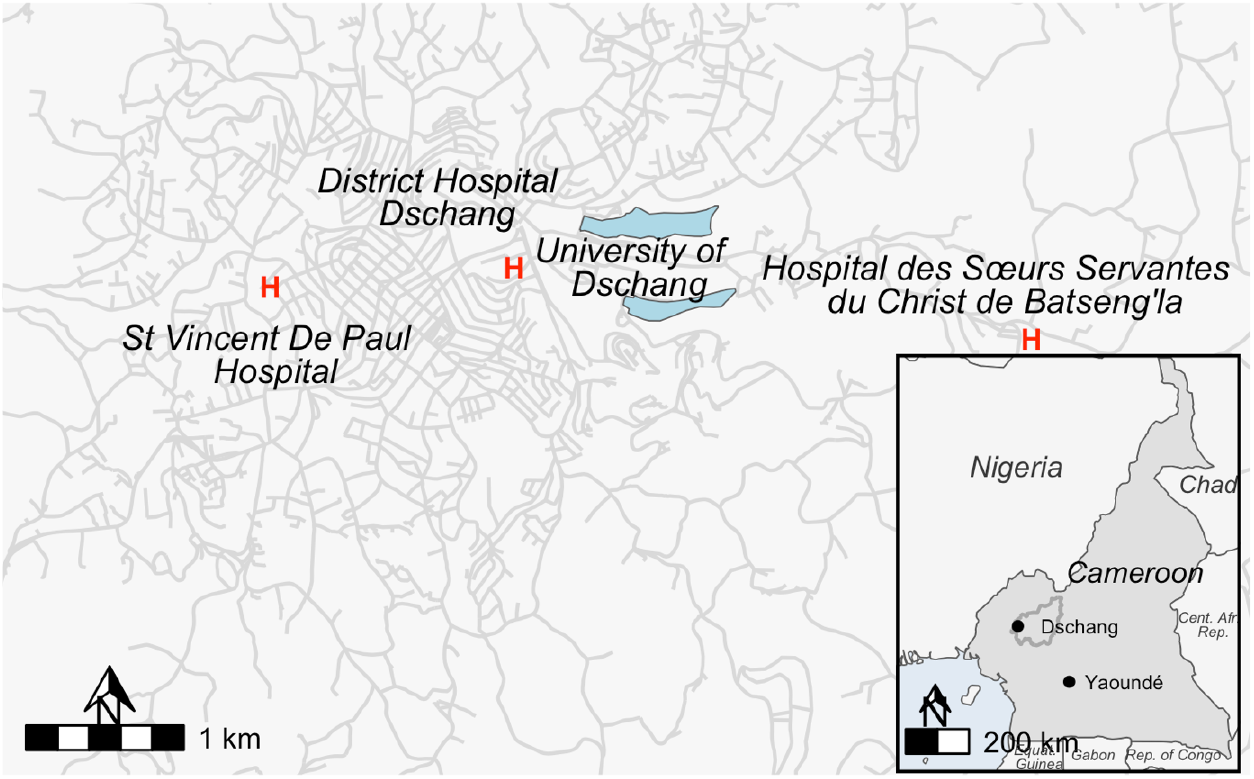
Clinical sites in Dschang, Cameroon.

**Figure 2:**
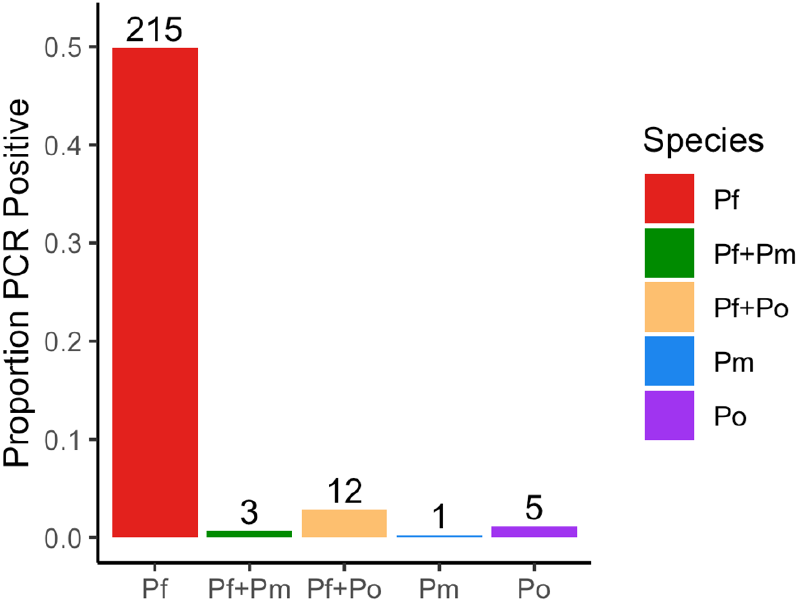
Molecular identification and distribution of *Plasmodium* spp. (*P. falciparum* vs. others)

Of the 17 *P. ovale* positive samples by real-time PCR, 12 were successfully classified to the species level. Six (35.3%) were *P. ovale curtisi (Poc)* and three (17.7%) were *P. ovale wallikeri (Pow)*. Mixed infections of *Pow/Poc* were identified in three (17.7%) samples. Although most (11/17; 64.7%) of these *P. ovale* positives were from Saint Vincent De Paul hospital, they were widely distributed across several quarters of Dschang. Moreover, 70.6% (12/17) were female. The most commonly reported symptoms amongst the *P. ovale* positives were headache 29.4% (5/17) followed by general fatigue 23.5% (4/17). Thirteen (76.5%) of these patients were identified to live within 1-km from lake sources. All the *P. ovale* mono infections came from 20- to 23-year-old patients with headache (3/5; 60 %) as this was the most represented symptom. No significant association was observed between *Plasmodium* species infections and clinical parameters and documented symptoms.

## Discussion

We investigated the prevalence of non-falciparum malaria in Dschang, West region, Cameroon using highly sensitive molecular testing among individual reporting to clinics with symptoms consistent with uncomplicated malaria. We showed that non-falciparum malaria is circulating within Dschang, resulting in clinical malaria cases, the majority of which are associated with mixed infections with *P. falciparum* and only a handful were due to non-falciparum mono-infections.

*P. falciparum* accounted for more than 90% of all malaria cases. This is consistent with other studies across Sub-Saharan Africa (3,26–28) and in Cameroon (29–32). Even though *P. falciparum* prefers young reticulocytes, it has the ability to invade all red blood cell (RBC) age classes, increasing the probability of having higher *P. falciparum* parasite density, thereby reducing RBC availability for other *Plasmodium* species (33). The low prevalence of *P. malariae* in this study does not align with other studies carried out in Cameroon. Indeed, Nguiffo-Nguete *et al*. (4) reported a high proportion of *P. malariae* infections in Adamaoua; reporting 2.5% *P. malariae* mono-infections as compared to 0.4% in our study and 17% *P. falciparum/P. malariae* co-infections compared to 1.3% in our study. A possible explanation could be the difference in transmission patterns in both epidemiological zones as well as the sample collection period (17,34). In these other studies, the authors looked at the transmission pattern of malaria species in several regions of Cameroon ranging from the sahelian region to the forest humid zone passing across the tropical area and the great highlands.

In this study, *P. vivax* was not identified, which contrasts with previous studies in Cameroon (8,12,37) and in Dschang in particular (6,7). The study conducted by Ngassa *et al*. (35) targeted 4 regions of Cameroon (Centre, Littoral, South, and East) and recruited symptomatic hospital cases. They reported at least one *P. vivax* infection either as a mono- or mixed infection with *P. falciparum* in all study sites. Triple infection with *P. malariae* as well as mono- and mixed infections was also reported by Fru-Cho *et al*. (12) in Bolifamba in the Southwest region of Cameroon. These reported levels of *P. vivax* are relatively low compared to levels reported by Russo *et al*. who sampled febrile patients attending the hospital (35% of all malaria infections in Dschang (West region of Cameroon)) (6). This suggested a potential *P. vivax* hotspot. Although we did not find *P. vivax* in our study, this may have been due to timing of sampling or other unmeasured variables. The previous reports support the need for wider sampling, not only of hospital cases, but also asymptomatic community cases.

Overall, we had a combined prevalence of 4% (17/431) for *P. ovale spp*. with 5 mono-infections and 12 co-infections with *P. falciparum*. We documented the presence and distribution of both species of *P. ovale* in Dschang either as single infections or in mixed infections. This study is consistent with reports from Kojom-Foko and colleagues (36) who were the first to report the presence of *Poc* in Douala, Cameroon (29). Our results corroborate findings from other studies conducted in countries neighboring Cameroon, such as Equatorial Guinea (37), the Republic of Congo (38) Nigeria and Gabon (39), which reported the presence of both sub-types. The detection of both *P. ovale* species in Dschang highlights the need to better describe the epidemiology and transmission of these species among populations in Cameroon. The relapsing malarias, *P. ovale spp*. and *P. vivax* deserve further study in Cameroon due to the need for understanding relapses and whether hypnozoite treatment with primaquine is needed. Equally important is the need to address burden of relapsing malaria in Cameroon due to their potential role in life-threatening malaria complications and severe thrombocytopenia (40). Together, these factors necessitate improved surveillance and diagnostic capabilities to accurately detect and differentiate the various species and subspecies of *Plasmodium* in Cameroon.

There are limitations to this study. We may have missed dormant *P. vivax* and *P. ovale* spp. infections due to their ability to form hypnozoites (41). Furthermore, the hospital-based approach for the collection of our samples limited us to clinical cases suspected for malaria infections, and non-falciparum infections may result in less subtle symptoms reducing the likelihood of seeking care, although previous studies of *P. vivax* from Dschang identified cases from symptomatic persons (6). Population-targeted sampling of asymptomatic individuals was not conducted, which may differ from symptomatic case prevalence estimates.

## Conclusion

We characterized the prevalence of Plasmodium species, particularly the non-falciparum species, among individuals presenting to clinic in Dschang in 2020. The results show that non-falciparum malaria species circulate in Dschang in non-negligible proportions, including both species of *Plasmodium ovale*. Non-falciparum malaria remains a concern locally in Dschang, and expanding community-level surveillance will help clarify its burden and transmission. In addition, much remains to be learned about *P. ovale curtisi* and *P. ovale wallikeri* biology and epidemiology in Sub-Saharan Africa, including the differential role they play in relapsing malaria, the dynamics of coinfections and with other species and transmission as well as how they may be evolving in against current malaria control efforts designed to target *P. falciparum*. Our findings suggest that the degree to which these closely related but sympatric species co-circulate within their human hosts may be under-appreciated. Further studies of non-falciparum malaria, in both febrile and asymptomatic individuals, across Cameroon will help us better understand these neglected pathogens and develop control measures to eliminate all malaria.

## Data Availability

All data produced in the present study are available upon reasonable request to the authors

## Author Contributions

Conceptualization: IMA, JJJ, JTL

Data curation: PVKT, VP

Formal analysis: PVKT, VP, EMA, DBNK, MSM

Funding acquisition: IMA, JTL

Investigation: PVKT, VP, OK

Methodology: IMA, JTL, JJJ

Project administration: PVKT, IMA, JJJ

Resources: IMA, JTL, RRD, JJJ

Supervision: IMA, JJJ

Validation: IMA, JTL, JJJ.

Writing – original draft: PVKT.

Writing – review & editing: IMA, SJW, VP, EMA, MSM, DBNK, OK, JP, JTL, CT, RRD, JJJ.

## Acknowledgments

We wish to acknowledge the administrative authorities of the Dschang Health District and the hospital personnel for providing the assistance with the field work. The following reagent was obtained through BEI Resources, NIAID, NIH: Diagnostic Plasmid Containing the Small Subunit Ribosomal RNA Gene (18S) from *Plasmodium falciparum*, MRA-177, contributed by Peter A. Zimmerman; Diagnostic Plasmid Containing the Small Subunit Ribosomal RNA Gene (18S) from *Plasmodium vivax*, MRA-178, contributed by Peter A. Zimmerman; Diagnostic Plasmid Containing the Small Subunit Ribosomal RNA Gene (18S) from *Plasmodium malariae*, MRA-179, contributed by Peter A. Zimmerman; Diagnostic Plasmid Containing the Small Subunit Ribosomal RNA Gene (18S) from *Plasmodium ovale*, MRA-180, contributed by Peter A. Zimmerman.

