## Supplemental Table 1 for "Distribution of non-falciparum malaria among symptomatic malaria patients in Dschang, West Region of Cameroon"

**Supplementary table S1: Primer sequences and reaction conditions**

^a^Hoffman, N, et. al. PLOS Medicine. 2015.

| **Target Gene** | **Primers** | **Primer Sequences** | **Number of cycles** | **Temperature (℃); time** | **Reaction condition** |
| --- | --- | --- | --- | --- | --- |
| ***Plasmodium falciparum* varATS^a^** | Forward | 5'- CCCATACACAACCAAYTGGA - 3' | 45 | 50; 2min  95;10min  95;15sec  55;1min | Fwd primer 800 nM Rev primer 800 nM Probe 400 nM Template DNA 2.5 µl Total volume 10 µl |
|  | Reverse | 5' - TTCGCACATATCTCTATGTCTATCT - 3' |  |  |  |
|  | Probe | 5' - 6-FAM-TRTTCCATAAATGGT-NFQ-MGB - 3' |  |  |  |
| ***Plasmodium ovale spp.* 18S rRNA^b^** | Forward | 5’ - CCRACTAGGTTTTGGATGAAAVRTTTTT- 3’ | 45 | 50; 2min 95;10min 95;15sec 52;1min | Fwd primer 400 nM Rev primer 400 nM Probe 200 nM Template DNA 4 µl Total volume 10 µl |
|  | Reverse | 5’ - AACCCAAAGACTTTGATTTCTCATAA - 3’ |  |  |  |
|  | Probe | 5’ - VIC/CRAAAGGAATTYTCTTATT - 3’ |  |  |  |
| ***Plasmodium vivax* 18S rRNA^c^** | Forward | 5’ - ACGCTTCTAGCTTAATCCACATAACT - 3’ | 45 | 50; 2min 95;10min 95;15sec 60;1min | Fwd primer 400 nM Rev primer 400 nM Probe 200 nM Template DNA 5 µl Total volume 10 µl |
|  | Reverse | 5’ - ATTTACTCAAAGTAACAAGGACTTCCAAGC - 3’ |  |  |  |
|  | Probe | 5' - /56-FAM/TTCGTATCG/ZEN/ACTTTGTGCGCATTTTGC/3IABkFQ/ - 3' |  |  |  |
| ***Plasmodium malariae* 18S rRNA^c^** | Forward | 5'- AGT TAA GGG AGT GAA GAC GAT CAG A - 3' | 45 | 50; 2min  95;10min 95;15sec 56;1min | Fwd primer 400 nM Rev primer 400 nM Probe 200 nM Template DNA 5 µl Total volume 10 µl |
|  | Reverse | 5'- CAA CCC AAA GAC TTT GAT TTC TCA TAA -3' |  |  |  |
|  | Probe | 5' - 6-FAM-ATG AGT GTT TCT TTT AGA TAG C-NFQ-M |  |  |  |

^b^Mitchell C, et. al. Journal of Infectious Diseases. 2021

^c^Brazeau N, et. al. Nature Communications. 2021
